# A systematic review of clinical health conditions predicted by machine learning diagnostic and prognostic models trained or validated using real-world primary health care data

**DOI:** 10.1101/2022.08.25.22279229

**Authors:** Hebatullah Abdulazeem, Sera Whitelaw, Gunther Schauberger, Stefanie J. Klug

**Author notes:** Corresponding author Dr. Hebatullah Abdulazeem Chair of Epidemiology Technical University of Munich Georg-Brauchle Ring 56 80993 Munich, Germany (HA).

## Abstract

**Aim:** With the rapid advances in technology and data science, machine learning (ML) is being adopted by the health care sector; but there is a lack of literature addressing the health conditions targeted by the ML prediction models within primary health care (PHC). To fill this gap in knowledge, we conducted a systematic review following the PRISMA guidelines to identify the health conditions targeted by ML in PHC.

**Methods:** We searched the Cochrane Library, Web of Science, PubMed, Elsevier, BioRxiv, Association of Computing Machinery (ACM), and IEEE Xplore databases for studies published from January 1990 to January 2022. We included any primary study addressing ML diagnostic or prognostic predictive models that were supplied completely or partially by real-world PHC data. We performed literature screening, data extraction, and risk of bias assessment. Health conditions were categorized according to international classification of diseases. Extracted date were analyzed quantitatively and qualitatively.

**Results:** We identified 109 studies investigating 42 health conditions. These studies included 273 ML prediction models supplied by the PHC data of 24.2 million participants from 19 countries. We found that 82% of the studies were retrospective. 76.6% of the studies reported diagnostic predictive ML models. 77% of all reported models aimed for models’ development without external validation. Risk of bias assessment revealed that 90.8% of the studies were of high or unclear risk of bias. The most frequently reported health conditions were Alzheimer’s disease and diabetes mellitus.

**Conclusions:** To the best of our knowledge, this is the first review to investigate the extent of the health conditions targeted by the ML prediction models within PHC settings. Our study provides an important summary on the presently available ML models in PHC, which can be used in further research and implementation efforts.

## Introduction

Primary health care (PHC) is considered the gatekeeper, where health education and promotion are provided, non-life-threatening health conditions are diagnosed and treated, and chronic diseases are managed [1]. This form of health maintenance, which aims to provide constant access to high-quality care and comprehensive services, is defined and called for by the WHO global vision for PHC [2]. To achieve these PHC care aims, common health disorders require risk prediction for primary prevention, early diagnosis, follow-up, and timely interventions to avoid diseases exacerbations and complications, all of which are the core practice of PHC [3].

With the high number of patients visiting PHC and the emerge of electronic health records, “Big Data” is generated with subsequent difficulties to be handled by traditional data analytics [4]. Tools that could more accurately predict diseases incidence and progression and offer advice on appropriate treatment could substantially improve the decision-making process. Machine Learning (ML), a subtype of artificial intelligence (AI), provides methods to productively mine this big data, such as predictive models that potentially forecast and predict diseases occurrence and progression [5].

Integrating the PHC medical efforts with the continuously updated technologies constitutes a fusion of numerous disciplines and views aimed at improving the performance of health care regarding patient care and the productivity and efficiency within health care facilities [5, 6]. ML models have been developed in health research – most significantly in the last decade - to predict the incidence of diabetes, cancers, and recently COVID-19 pandemic related illness from health records [7]. A systematic overview of 35 studies published in 2021 investigated the existing literature of AI/ML, but exclusively in relation to World Health Organization indicators [8]. Other literature and scoping reviews examined AI/ML in relation to certain health conditions, such as HIV [9], hypertension [10], and diabetes [11]. Other systematic reviews targeted specific health conditions across multiple health sectors, such as pregnancy care [12], melanoma [13], stroke [14], and diabetes [15]. However, reviews investigating PHC specifically have been fewer [16, 17]. It has been reported that research on ML for PHC stands at an early stage of maturity [17]. Similar to ours, a recently published protocol of a systematic review addressing the performance of ML prediction models in multiple different medical fields was published [18]. However, this protocol does not focus on primary care in specific and its search is limited to the years 2018 and 2019. Hence, the current literature is not enough to identify what the diseases targeted by ML prediction models within the real-world primary care are. Furthermore, literature investigating validity and potential impact on health of such models are not abundant. To address this gap, our objective was to encompass the health conditions predicted by using ML models to identify and assess the extent of the body of research within real-world PHC settings.

## Methods

We conducted a systematic review in accordance with the Preferred Reporting Items for Systematic Reviews and Meta-Analyses (PRISMA) [19] and the CHecklist for critical Appraisal and data extraction for systematic Reviews of prediction Modelling Studies (CHARMS) [20]. The protocol for our review was registered on PROSPERO CRD42021264582 [21].

We included primary research articles (peer-reviewed, preprint, or abstract) published in any language. Studies that were published between January 1, 1990, when ML algorithm with a data-driven approach was first developed [22] and January 4, 2022 were included. Studies that reported real-world exclusive or mixed PHC data for any health condition in ambulatory settings, including referred patients from PHC to other health care facilities, worldwide were included. Studies that reported any prediction ML models within the PHC level that was classified as AI, DL or ML models were included.

### Search strategy and selection criteria

A comprehensive and systematic search was performed covering multidisciplinary databases: 1. Cochrane Library, 2. Elsevier (including ScienceDirect, Scopus, and Embase), 3. PubMed, 4. Web of Science (including nine databases), 5. BioRxiv and MedRxiv, 6. Association for Computer Machinery (ACM) Digital Library, and 7. Institute of Electrical and Electronics Engineers (IEEE) Xplore Digital Library.

To identify potentially relevant studies, we searched literature with the last updated search on January 4, 2022, back to January 1, 1990. The utilized search terms included “machine learning”, “artificial intelligence”, “deep learning”, and “primary health care”. Boolean operators and symbols were adapted to each literature database. Hand searches of citations of relevant reviews and a cross-reference check of the retrieved articles was also performed. Conference abstracts and gray literature searches were conducted using the available features of some databases. The full search strategy for all the electronic databases is presented in S1 Appendix. A reference management software (EndNote X9) was used to import references and to remove duplicates.

### Literature screening, data collection and statistical analysis

Title and abstract screening for all records were conducted independently by two researchers through the Rayyan platform [23]. Discrepancies were resolved by discussion. All studies that met the eligibility criteria were included in the systematic review.

The data extracted included: meta-data (first author, year, and publisher), source of primary data (country under investigation), datasets used (exclusive PHC data that was generated only within PHC settings or mixed data that was generated within PHC settings in addition to other data sources, such as secondary or tertiary health care), period of data extraction, sample size, and study design, predicted health condition, study objectives (incident diagnostic, prevalent diagnostic or prognostic), aim of model proposal (development without external validation, development with external validation, or external validation without or with update). Data extraction was performed by two authors.

Health conditions extracted were categorized according to international classification of diseases (ICD)-10 version 2019 [24]. Further categorization was based on the ML models’ aim. Descriptive statistics (number and percentage of studies) were calculated. Additionally, the overall number of participants was calculated, taking into consideration the potential overlap between the included datasets. This overlap assessment was identified based on similarity of datasets, period of data gathering within each included study, the targeted health condition and the inclusion and exclusion criteria of the participants. The quantitative results were calculated using Microsoft Excel.

### Risk of bias and applicability assessment

The ‘Prediction model study Risk Of Bias Assessment Tool’ (PROBAST) was used to assess the risk of bias and concerns about the applicability of the included studies [25]. The four domains of this tool, which are participants, predictors, outcome, and analysis were addressed. The overall judgement for the risk of bias evaluation and concern of applicability of the prediction models in PROBAST is ‘low,’ ‘high,’ or ‘unclear.’ In cases when all domains were graded ‘low’ risk of bias, assessment of ‘models developed without external validation’ was downgraded to ‘high’ risk of bias even if all the four domains were of low risk of bias, unless the model’s development was based on an exceptionally large sample size and included some form of internal validation. Results of risk of bias and concern of applicability assessments were presented in a color-coded graph.

## Results

Our search strategy yielded 23,045 publications. After duplicate removal, 19,280 publications were screened, of those 167 publications were eligible for the full text screening. A total of 109 publications met our inclusion criteria (Fig 1). A list of the excluded studies with the justification of exclusion is presented in S2 Appendix.

**Figure 1.**
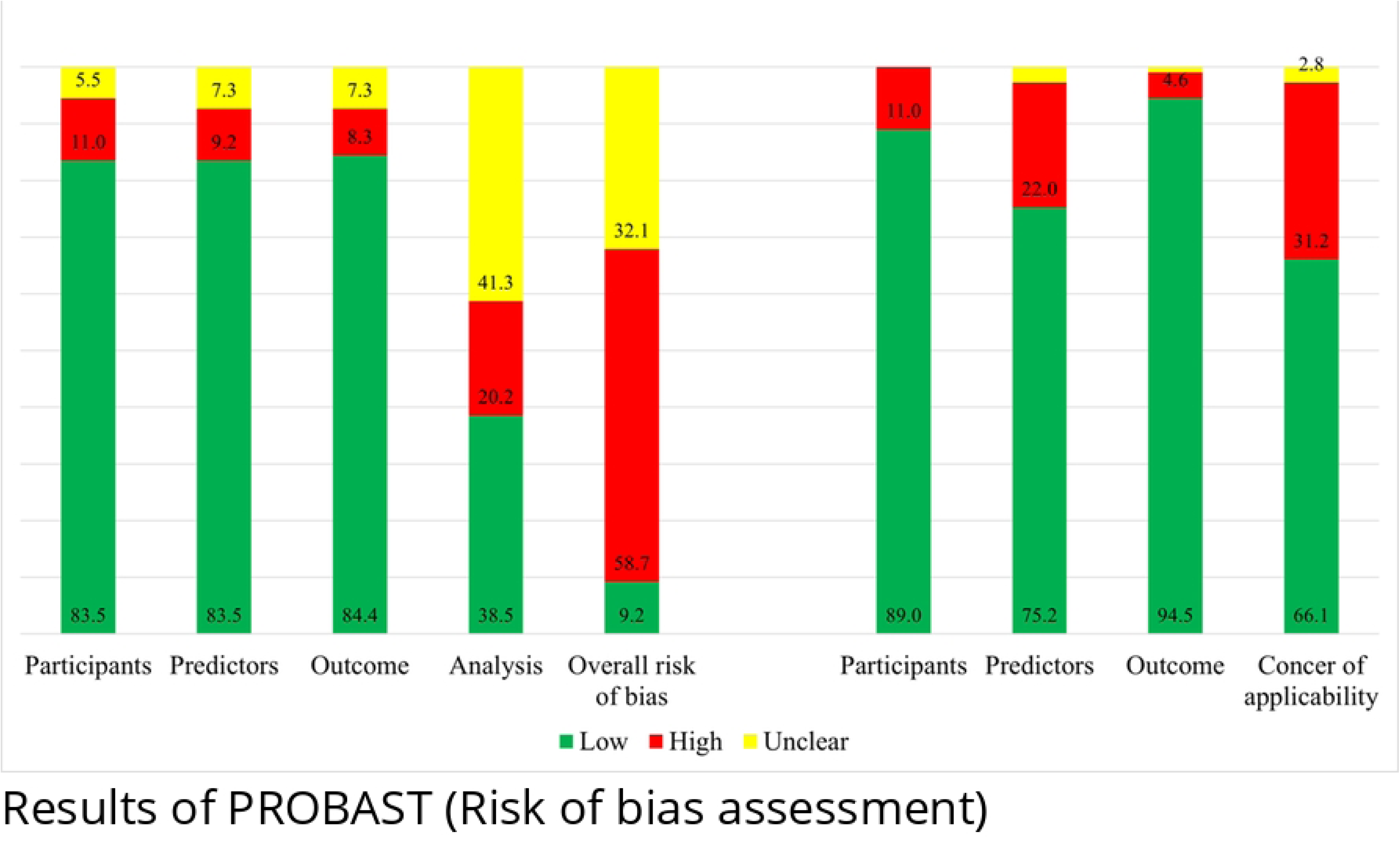
Prisma Flow diagram

The results of the data extracted in this review are presented in the following paragraphs in the form of geographical and chronological characteristics, studies’ design and the ML models addressed, and the health conditions investigated. Additionally, three tables, Table 1, 2, and 3, are depicting the characteristics of the included studies. Table 1 presents the studies that reported only developing ML prediction models without implication of any external validation of the models developed. Table 2 presents the studies that reported both developing and validating ML prediction models. Whereas Table 3 presents the studies that reported only the validation of previously developed models. In both Table 2 and 3, each row represents different dataset that was used to develop and/or validate the prediction models.

**Table 1.** Overview of the included studies reporting ML prediction models developed using primary health care data without conducting external validation (n = 84)

| Study | Study design | Objective | Country | Source of data set | Population | Period of extracted data | Health condition |
| --- | --- | --- | --- | --- | --- | --- | --- |
| **Circulatory System Diseases** |  |  |  |  |  |  |  |
| Chen et al (2019) [46] | Retro. nested case control | Incident diagnostic | United States | Exclusive | 34,502 | 05/2000-05/2013 | Heart failure |
| Choi et al (2017) [47] | Retro. case control | Incident diagnostic | United States | Exclusive | 32,787 | 05/2000-05/2013 | Heart failure |
| Du et al (2020) [48] | Retro. cohort | Prognostic | China | Linked | 42,676 | 2010-2018 | Hypertension |
| Farran et al (2013) [49] | Retro. cohort | Incident diagnostic | Kuwait | Linked | 270,172 | 12 years | Any cardiovascular disease |
| Hill et al (2019) [37] | Retro. cohort | Incident diagnostic | United Kingdom | Exclusive | 2,994,837 | 01/2006-12/2016 | Atrial fibrillation |
| Karapetyan et al (2021) [50] | Retro. cohort | Prognostic | Germany | Exclusive | 46,071 | 02-2020-09/2020 | Any cardiovascular disease |
| Lafreniere et al (2017) [51] | Retro. cohort | Incident diagnostic | Canada | Exclusive | 379,027 | Not reported | Hypertension |
| Li et al (2020) [43] | Retro. cohort | Incident diagnostic | United Kingdom | Linked | 3,661,932 | 01/1998-12/2018 | Any cardiovascular disease |
| Lip et al (2021) [52] | Retro. cohort | Prevalent diagnostic | Australia | Linked | 926 | Not reported | Hypertension |
| Lorenzoni et al (2019) [53] | Pros. cohort | Prognostic | Italy | Linked | 380 | 2011-2015 | Heart failure |
| Ng et al (2016) [54] | Retro. nested case control | Incident diagnostic | United States | Exclusive | 152,095 | 2003-2010 | Heart failure |
| Nikolaou et al (2021) [55] | Pros. cohort | Prognostic | United Kingdom | Exclusive | 6,883 | 2015-2019 | Any cardiovascular disease |
| Saraju et al (2021) [56] | Retro. cohort | Incident diagnostic | United States | Exclusive | 32,192 | 01/2009-12/2018 | Any cardiovascular disease |
| Selskyy et al (2018) [57] | Retro. case control | Prognostic | Ukraine | Exclusive | 63 | 2011-2012 | Hypertension |
| Shah et al (2019) [27] | Retro. cohort | Prognostic/ Incident diagnostic | United Kingdom | Linked | 2,000 | Not reported | Myocardial infarction |
| Solanki et al (2020) [58] | Retro. cohort | Prevalent diagnostic | United States | Exclusive | 495 | 2007-2017 | Hypertension |
| Solares et al (2019) [59] | Pros. cohort | Incident diagnostic | United Kingdom | Exclusive | 80,964 | entry – 01/2014 | Any cardiovascular disease |
| Ward et al (2020) [60] | Retro. cohort | Incident diagnostic | United States | Exclusive | 262,923 | 01/2009-12/2018 | Atherosclerosis |
| Weng et al (2017) [35] | Pros. cohort | Incident diagnostic | United Kingdom | Exclusive | 378,256 | 01/2005-01/2015 | Any cardiovascular disease |
| Wu et al (2010) [61] | Retro. case control | Incident diagnostic | United States | Exclusive | 44,895 | 01/2003-12/2006 | Heart failure |
| Zhao et al (2020) [34] | Retro. cohort | Incident diagnostic | United States | Linked | 4,914 | Not reported | Stroke |
| **Digestive System Diseases** |  |  |  |  |  |  |  |
| Sáenz Bajo et al (2002) [36] | Retro. cohort | Prevalent diagnostic | Spain | Exclusive | 81 | 01/1999-06/1999 | Gastroesophageal reflux |
| Waljee et al (2018) [62] | Retro. cohort | Prognostic | United States | Linked | 20,368 | 2002-2009 | Inflammatory bowel disorders |
| **Endocrine, Metabolic, and Nutritional Diseases** |  |  |  |  |  |  |  |
| Akyea et al (2020) [63] | Retro. cohort | Incident diagnostic | United Kingdom | Exclusive | 4,027,775 | 01/1999-06/2019 | Familial hypercholesterolemia |
| Álvarez-Guisasaola et al (2010) [42] | Pros. cohort | Incident diagnostic | Spain | Exclusive | 2,662 | Not reported | Diabetes mellitus |
| Crutzen et al (2021) [64] | Retro. cohort | Incident diagnostic | The Netherlands | Exclusive | 138,767 | 01/2007-01/2014 | Diabetes mellitus |
| Ding et al (2019) [40] | Retro. case control | Prevalent diagnostic | United States | Exclusive | 97,584 | 1997-2017 | Primary Aldosteronism |
| Dugan et al (2015) [38] | Retro. cohort | Prognostic | United States | Exclusive | 7,519 | Over 9 years | Obesity |
| Farran et al (2019) [65] | Retro. cohort | Prognostic | Kuwait | Exclusive | 1,837 | Over 9 years | Diabetes mellitus |
| Hammond et al (2019) [66] | Retro. cohort | Prognostic | United States | Exclusive | 3,449 | 01/2008-08/2016 | Obesity |
| Kopitar et al (2020) [67] | Retro. case control | Incident diagnostic | Slovenia | Exclusive | 27,050 | 12/2014-09/2017 | Diabetes mellitus |
| Lethebe et al (2019) [68] | Retro. cohort | Prevalent diagnostic | Canada | Exclusive | 1,309 | 2008-2016 | Diabetes mellitus |
| Looker et al (2015) [69] | Pros. nested case control | Prognostic | United Kingdom | Exclusive | 309 | 12/1998-05/2009 | Diabetic nephropathy |
| Metsker et al (2020) [70] | Retro. cohort | Incident diagnostic | Russia | NR | 54,252 | 07/2009-08/2017 | Diabetic polyneuropathy |
| Metzker et al (2020) [71] | Retro. cohort | Incident diagnostic | Russia | NR | 58,462 | Not reported | Diabetic polyneuropathy |
| Nagaraj et al (2019) [72] | Retro. cohort | Prognostic | The Netherlands | Exclusive | 11,887 | 01/2007-12/2013 | Diabetes mellitus |
| Pakhomov et al (2008) [31] | Retro. cohort | Prevalent diagnostic | United States | Exclusive | 145 | 07/2004-09/2004 | Diabetic foot |
| Rumora et al (2021) [73] | Cross sectional | Incident diagnostic | Denmark | Exclusive | 97 | 10/2015-06/2016 | Diabetic polyneuropathy |
| Tseng et al (2021) [33] | Cross sectional | Incident diagnostic | United States | Exclusive | NR | Not reported | Diabetes mellitus |
| Wang et al (2021) [74] | Retro. cohort | Incident diagnostic | China | Exclusive | 1,139 | 2017-2019 | Gestational diabetes |
| Willimson et al (2020) [75] | Pros. cohort | Incident diagnostic | United States | Linked | 866 | Not reported | Familial hypercholesterolemia |
| **External Cause of Mortality** |  |  |  |  |  |  |  |
| DelPozo-Banos et al (2018) [76] | Retro. case control | Incident diagnostic | United Kingdom | Linked | 54,684 | 2001-2015 | Suicidality |
| Penfold et al (2021) [77] | Retro. cohort | Incident diagnostic | United States | Linked | 256,823 | Not reported | Suicide |
| van Mens et al (2020) [78] | Retro. case control | Incident diagnostic | The Netherlands | Exclusive | 207,882 | 2017 | Suicidality |
| **Genitourinary System Diseases** |  |  |  |  |  |  |  |
| Shih et al (2020) [79] | Retro. cohort | Incident diagnostic | Taiwan | Linked | 19,270 | 01/2015-12/2019 | Chronic kidney disease |
| Zhao et al (2019) [80] | Pros. cohort | Incident diagnostic | United States | Exclusive | 61,740 | 2009–2017 | Chronic kidney disease |
| **Mental and Behavioral Diseases** |  |  |  |  |  |  |  |
| Alexander et al (2021) [28] | Retro. cohort | Prevalent diagnostic | United Kingdom | Exclusive | 7,913 | 01/1997-06/2016 | Alzheimer's disease |
| Dinga et al (2018) [81] | Pros. cohort | Prognostic | The Netherlands | Linked | 804 | Not reported | Major depressive disorder |
| Ford et al (2019) [82] | Retro. case control | Incident diagnostic | United Kingdom | Exclusive | 93,120 | 2000-2012 | Alzheimer's disease |
| Ford et al (2020) [83] | Retro. case control | Incident diagnostic | United Kingdom | Exclusive | 95,202 | 2000-2012 | Alzheimer's disease |
| Ford et al (2021) [84] | Retro. case control | Prevalent diagnostic | United Kingdom | Exclusive | 93,426 | 2000-2012 | Alzheimer's disease |
| Fouladvand et al (2019) [85] | Retro. cohort | Prognostic | United States | Exclusive | 3,265 | Not reported | Alzheimer's disease |
| Haun et al (2021) [86] | Cross sectional | Incident diagnostic | Germany | Exclusive | 496 | Not reported | Anxiety |
| Jammeh et al (2018) [87] | Retro. Case control | Incident diagnostic | United Kingdom | Exclusive | 3,063 | 06/2010-06/2012 | Alzheimer's disease |
| Jin et al (2019) [88] | Retro. Cohort | Incident diagnostic | United States | Exclusive | 923 | 2010-2013 | Major depressive disorder |
| Kaczmarek et al (2019) [89] | Retro. Case control | Prevalent diagnostic | Canada | Exclusive | 890 | Not reported | Post-traumatic stress disorder |
| Ljubic et al (2020) [90] | Retro. Cohort | Incident diagnostic | United States | Exclusive | 2,324 | Not reported | Alzheimer's disease |
| Mallo et al (2020) [91] | Retro. case control | Prognostic | Spain | Exclusive | 128 | 2008 | Alzheimer's disease |
| Mar et al (2020) [92] | Retro. case control | Prevalent diagnostic | Spain | Linked | 4,003 | Not reported | Alzheimer's disease |
| Pólchlopek et al (2020) [93] | Retro. cohort | Incident diagnostic | The Netherlands | Exclusive | 92,621 | 2007-12/2016 | Any mental disorder |
| Shen et al (2020) [94] | Retro. cohort | Incident diagnostic | China | Exclusive | 2,299 | 2008-2018 | Alzheimer's disease |
| Suárez-Araujo et al (2021) [95] | Retro. case control | Prevalent diagnostic | United States | Exclusive | 330 | Not reported | Alzheimer's disease |
| Tsang et al (2021) [96] | Retro. cohort | Prognostic | United Kingdom | Exclusive | 592,988 | 1982-2015 | Alzheimer's disease |
| Zafari et al (2021) [97] | Retro. cohort | Incident diagnostic | Canada | Exclusive | 154,118 | 01/1995-12/2017 | Post-traumatic stress disorder |
| **Musculoskeletal and Connective Tissue Diseases** |  |  |  |  |  |  |  |
| Emir et al (2014) [98] | Retro. cohort | Incident diagnostic | United States | Exclusive | 587,961 | 2011-2012 | Fibromyalgia |
| Javrik et al (2018) [99] | Pros. cohort | Prognostic | United States | Exclusive | 3,971 | 03/2011-03/2013 | Back pain |
| Kennedy et al (2021) [100] | Retro. case control | Incident diagnostic | United Kingdom | Linked | 23,528 | Over 6 years | Ankylosing spondylitis |
| **Neoplasms** |  |  |  |  |  |  |  |
| Kop et al (2016) [101] | Retro. cohort | Incident diagnostic | The Netherlands | Exclusive | 260,000 | Not reported | Colorectal cancer |
| Malhotra et al (2021) [102] | Pros. case control | Incident diagnostic | United Kingdom | Exclusive | 56,953 | 01/2005-06/2009 | Pancreatic cancer |
| Ristanoski et al (2021) [103] | Retro. case control | Incident diagnostic | Australia | Exclusive | 683 | 2016-2017 | Lung cancer |
| **Nervous System Diseases** |  |  |  |  |  |  |  |
| Cox et al (2016) [104] | Retro. case control | Prevalent diagnostic | United Kingdom | Exclusive | 35,510 | 01/2007-12/2011 | Post stroke spasticity |
| Hrabok et al (2021) [105] | Retro. cohort | Prognostic | United Kingdom | Exclusive | 10,499 | 01/2000-05/2012 | Epilepsy |
| Kwasny et al (2021) [106] | Retro. case control | Incident diagnostic | Germany | Exclusive | 3,274 | 01/2010-12/2017 | Progressive supranuclear palsy |
| **Respiratory System Diseases** |  |  |  |  |  |  |  |
| Afzal et al (2013) [107] | Retro. cohort | Prevalent diagnostic | The Netherlands | Exclusive | 5,032 | 01/2000-01/2012 | Asthma |
| Doyle et al (2020) [108] | Retro. case control | Incident diagnostic | United Kingdom | Exclusive | 112,784 | 09/2003-09/2017 | Nontuberculous mycobacterial lung disease |
| Kaplan et al (2020) [109] | Retro. cohort | Prevalent diagnostic | United States | Linked | 411,563 | Not reported | Asthma/obstructive pulmonary disease overlap |
| Lisspers et al (2021) [110] | Retro. cohort | Prognostic | Sweden | Linked | 29,396 | 01/2000-12/2013 | Asthma |
| Marin-Gomez et al (2021) [111] | Retro. cohort | Incident diagnostic | Spain | Exclusive | 7,314 | 03/04/2020 | COVID-19 |
| Nikolaou et al (2021) [112] | Pros. cohort | Prognostic | United Kingdom | Exclusive | 13,260 | 2015-2018 | Chronic Obstructive Pulmonary Disease |
| Pikoula et al (2019) [30] | Retro. cohort | Prognostic | United Kingdom | Linked | 30,961 | 01/1998-01/2016 | Chronic Obstructive Pulmonary Disease |
| Stållberg et al (2021) [113] | Retro. cohort | Prognostic | Sweden | Linked | 7,823 | 01/2000-12/2013 | Chronic Obstructive Pulmonary Disease |
| Stephens et al (2020) [32] | Retro. case control | Incident diagnostic | United States | Exclusive | 7,278 | 2009-2019 | Influenza |
| Trtica-Majnaric et al (2010) [114] | Retro. cohort | Prognostic | Croatia | Exclusive | 90 | 2003-2004 | Influenza |
| Zafari et al (2022) [115] | Retro. cohort | Incident diagnostic | Canada | Exclusive | NR | Not reported | Chronic Obstructive Pulmonary Disease |

**Table 2.** Overview of the included studies reporting ML prediction models developed using primary health care data with conduction of external validation using different datasets (n = 13)

| Study | Study design | Objective | Country | Source of dataset | Population | Period of extracted data | Health condition |
| --- | --- | --- | --- | --- | --- | --- | --- |
| Endocrine, Metabolic, and Nutritional Diseases |  |  |  |  |  |  |  |
| Herttroijs et al (2018) [116] | Retro. cohort | Prognostic | Netherlands | Exclusive | 10,528 | 01/2006-12/2014 | Diabetes mellitus |
|  |  |  | Netherlands | Exclusive | 3,337 | 01/2009-12/2013 |  |
| Myers et al (2019) [117] | Retro. case control | Incident diagnostic | United States | Exclusive | 33,086 | 09/2013-08/2016 | Familial hypercholesterolemia |
|  |  |  | United States | Linked | 7,805 |  |  |
|  |  |  | United States | Linked | 35,090 |  |  |
|  |  |  | United States | Linked | 8,094 |  |  |
| Perveen et al (2019) [29] | Retro. cohort | Prognostic | Canada | Exclusive | 911 | 08/2003-06/2015 | Diabetes mellitus |
|  |  |  | Canada | Exclusive | 1,970 |  |  |
| Weisman et al (2020) [118] | Retro. cohort | Prevalent diagnostic | Canada | Exclusive | 5,402 | 2010-2017 | Diabetes mellitus |
|  |  |  | Canada | Linked | 29,371 |  |  |
| Mental and Behavioral Diseases |  |  |  |  |  |  |  |
| Amit et al (2021) [119] | Retro. cohort | Prevalent diagnostic | United Kingdom | Exclusive | 24,612 | 2000-2010 | Post-partum depression |
|  |  |  | United Kingdom | Exclusive | 9,193 | 2010-2017 |  |
|  |  |  | United Kingdom | Exclusive | 34,525 | 2000-2017 |  |
| Levy et al (2018) [120] | Pros. cohort | Incident diagnostic | United States | Exclusive | 49 | Over 9 months | Alzheimer’s disease |
|  |  |  | United States | Linked | 26 | Not reported |  |
| Perlis (2013) [121] | Retro. cohort | Prognostic | United States | Exclusive | 2,094 | 1999-2006 | Major depressive disorder |
|  |  |  | United States | Exclusive | 461 |  |  |
| Raket et al (2020) [45] | Retro. case control | Incident diagnostic | United States | Exclusive | 145,720 | 1990-2018 | Psychosis |
|  |  |  | United States | Exclusive | 4,770 |  |  |
| Musculoskeletal and Connective Tissue Diseases |  |  |  |  |  |  |  |
| Fernandez-Gutierrez et al (2021) [122] | Retro. cohort | Incident diagnostic | United Kingdom | Linked | 19,314 | 2002-2012 | Rheumatoid arthritis & Ankylosing spondylitis |
|  |  |  | United Kingdom | Linked | 1,868 |  |  |
| Jorge et al (2019) [123] | Retro. cohort | Incident diagnostic | United States | Linked | 400 | Not reported | Systematic lupus erythematosus |
|  |  |  | United States | Linked | 173 | Not reported |  |
| Zhou et al (2017) [124] | Retro. cohort | Incident diagnostic | United Kingdom | Linked | Unclear | 10/2013-07/2014 | Rheumatoid arthritis |
|  |  |  | United Kingdom | Linked | 475,580 | 03/2009-10/2012 |  |
| Neoplasms |  |  |  |  |  |  |  |
| Kinar et al (2016) [125] | Retro. cohort | Incident diagnostic | Israel | Exclusive | 606,403 | 01/2003-07/2011 | Colorectal cancer |
|  |  |  | United Kingdom | Exclusive | 30,674 | 01/2003-05/2012 |  |
| Pregnancy, Childbirth, Puerperium |  |  |  |  |  |  |  |
| Sufriyana et al (2020) [126] | Retro. nested case control | Incident diagnostic | Indonesia | Linked | 20,975 | 2015-2016 | Preeclampsia |
|  |  |  | Indonesia | Linked | 1,322 | Not reported |  |
|  |  |  | Indonesia | Linked | 904 | Not reported |  |

**Table 3.** Overview of the included studies reporting previously developed ML prediction models conducting only external validation using primary health care data (n = 12)

| Study | Study design | Objective | Country | Source of dataset | Population | Period of extracted data | Health condition |
| --- | --- | --- | --- | --- | --- | --- | --- |
| **Circulatory System Diseases** |  |  |  |  |  |  |  |
| Kostev et al (2021) [127] | Retro. cohort | Incident diagnostic | Germany | Exclusive | 11,466 | 01/2010-12/2018 | Stroke |
| Sekelj et al (2020) [128] | Retro. cohort | Incident diagnostic | United Kingdom | Exclusive | 604,135 | 01/2001-12/2016 | Atrial fibrillation |
| **Endocrine, Metabolic, and Nutritional Diseases** |  |  |  |  |  |  |  |
| Abramoff et al (2019) [129] | Pros. cohort | Prevalent diagnostic | United States | Exclusive | 819 | 01/2017-07/2017 | Diabetic retinopathy |
| Bhaskaranand et al (2019) [130] | Retro. cohort | Prevalent diagnostic | United States | Exclusive | 1,017,001 | 01/2014-09/2015 | Diabetic retinopathy |
| González-Gonzalo et al (2019) [131] | Retro. case control | Prevalent diagnostic | Spain | Exclusive | 288 | 08/2011-10/2016 | Diabetic retinopathy |
|  |  |  | Sweden | Exclusive |  |  |  |
|  |  |  | Denmark | Exclusive |  |  |  |
|  |  |  | United States | Linked | 4,613 | Over 2014 |  |
|  |  |  | United Kingdom | Linked |  |  |  |
| Kanagasingam et al (2018) [132] | Pros. cohort | Incident diagnostic | Australia | Exclusive | 193 | 12.2016-05/2017 | Diabetic retinopathy |
| Verbraak et al (2019) [133] | Retro. cohort | Prevalent diagnostic | The Netherlands | Exclusive | 1,425 | 2015 | Diabetic retinopathy |
| Neoplasms |  |  |  |  |  |  |  |
| Birks et al (2017) [134] | Retro. case control | Incident diagnostic | United Kingdom | Exclusive | 2,550,119 | 01/2000-04/2015 | Colorectal cancer |
| Hoogendoorn et al (2016) [135] | Retro. case control | Prevalent diagnostic | The Netherlands | Exclusive | 90,000 | 07/2006-12/2011 | Colorectal cancer |
| Hornbrook et al (2017) [136] | Retro. case control | Incident diagnostic | United States | Linked | 17,095 | 1998-2013 | Colorectal cancer |
| Kinar et al (2017) [137] | Pros. cohort | Incident diagnostic | Israel | Linked | 112,584 | 07/2007-12/2007 | Colorectal cancer |
| Respiratory System Diseases |  |  |  |  |  |  |  |
| Morales et al (2018) [41] | Retro. cohort | Prognostic | United Kingdom | Exclusive | 2,044,733 | 01/2000-04/2014 | Chronic Obstructive Pulmonary Disease |

### Geographical and chronological characteristics

The earliest included study was published in 2002, with the most publications occurring over the past four years. 77.9% (n= 85/109) of the publications were published between 2018 – 2021, (Fig 2). The United States of America (US) and the United Kingdom (UK) were reported in 58.1% of the included publications. While the 109 included publications reported countries 129 times, the US was reported 41 times and the UK 34 times. Other countries were identified but less frequently as depicted in S3 Figure. Usage of exclusive real-world primary health care data as predictors was reported in 77.4% (117 of 151 counts of data sources) across the studies. The remaining 22.6% of the PHC data sources were linked to different data sources, such as health insurance claims, cancer registries, secondary or tertiary health care, or administrative data. In the US, data was obtained mainly from PHC centers. In contrast, the most common source of the UK data was the Clinical Practice Research Datalink (CPRD), which is the largest patients’ data registry in the UK [26]. The period of data collection through the studies ranged from 1982 to 2020. The timeframe of patients’ data extracted and used to develop and train the ML models among the included studies varied between 2 months and 28 years. Sample sizes used for training and/or validating the models across the included studies ranged from 26 to around 4 million participants. A total number of participants within all the included studies was of 24.2 million. The potential overlaps of the datasets through each publication were investigated using two criteria, which were periods of the data extracted and participant characteristics per study. After identifying the potential overlap, the total number of unique participants was estimated to be 23.7 million.

**Figure 2.**
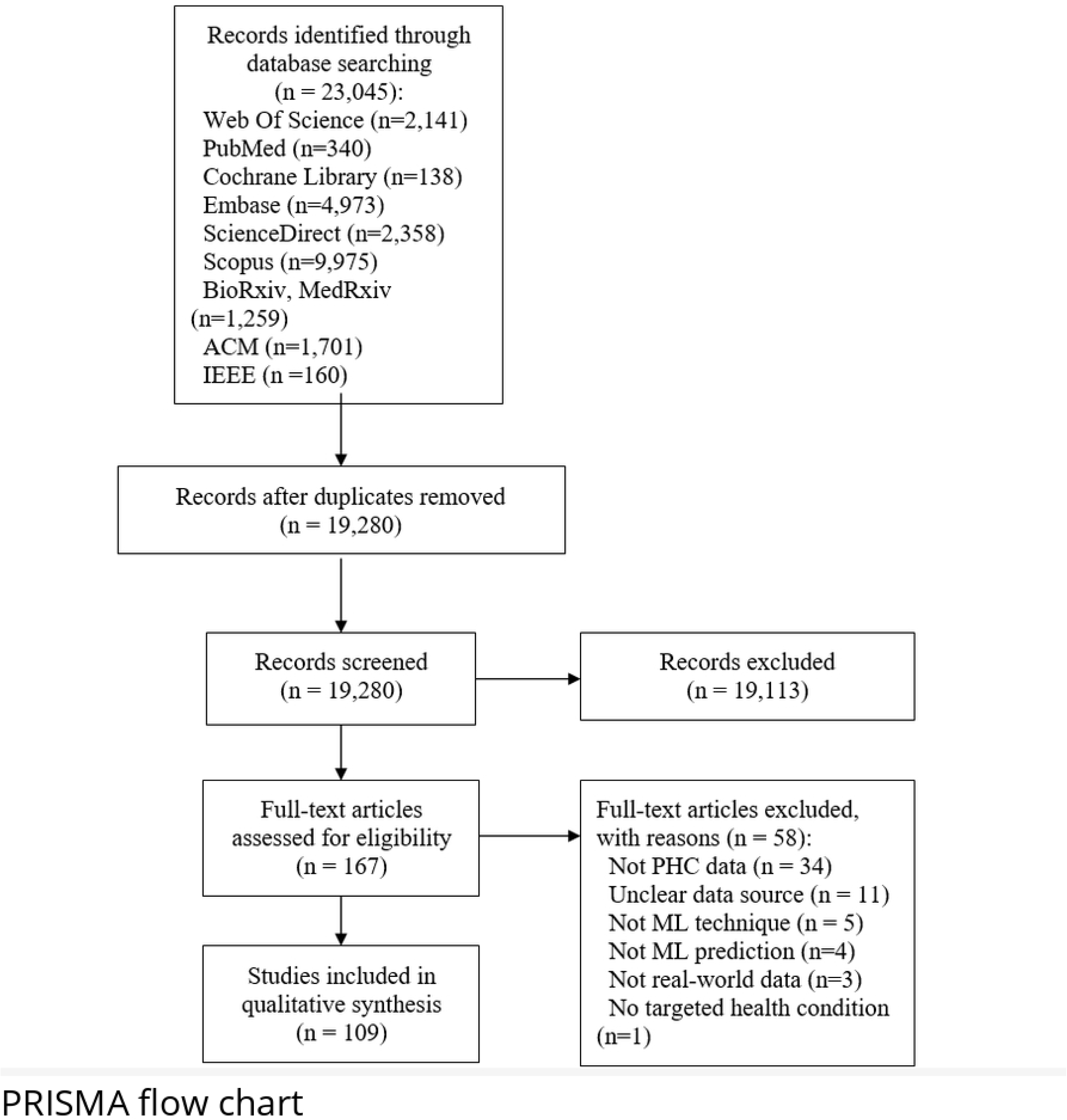
Number of studies per year of publication until December 31, 2021, in addition to one study included up to January 4, 2022.

### Studies’ design, objectives, and models

All the included studies were observational in design. Apart from 16 prospective studies, 85.3% (n= 93 of 109) of the studies were retrospective in design, of which 60 studies were reported as retrospective cohorts. The other reported studies designs were depicted in the supplementary S5 Figure. Regarding the primary objective of the included studies, 76.6% (n= 83 of 109) of the studies were predicting diagnosis of health conditions, either incident (n= 62 of 83) or prevalent (n= 21 of 83). The remaining 23.8% (n= 27 of 109, including one study of two different objectives [27]) predicted prognosis of health conditions, such as remission, improvement, complications, hospitalization, or mortality.

According to CHARMS guidelines, as mentioned earlier, the aim of the studies to use the prediction models can be one of three aims. These aims are model development without external validation, model development with external validation, and external validation of a predeveloped model with or without further model update [20]. The main aim of the included studies was found to be development of prediction models without evaluating the generalizability of the models, i.e., external validations (77%, n= 84 of 109). Another 13 (11.9%) studies developed and externally validated the models and only 12 studies (11%) externally validated previously existing models, but none of these studies reported updating the assessed model.

Within the 109 included publications, 273 models were developed and/or validated. The most frequent used type of ML was the supervised learning 84.2% (n= 230 of 273 models across the included studies). These supervised ML models were identified as follows: random forest (n= 53), logistic regression (n= 42), support vector machine (n= 33), boosting models such as extreme, light, and adaptive boosting (n= 29), decision tree (n= 28), and others such as naïve bias, k-nearest neighbors, and Least Absolute Shrinkage and Selection Operator (LASSO) (n= 45). Reinforcement ML/deep learning techniques, such as neural networks, were reported 36 times (13.1%, of 273 models), cross the studies, either exclusively or in comparison to other supervised ML models. A few studies (n= 3 of 109 studies) developed seven unsupervised ML models, such as k-means for predicting diseases prognosis through clustering it with the other morbidities [28–30]. A few studies (n= 5) used the natural language processing (NLP) technique as a preparatory step for using the free text clinical notes as (additional) predictors [27,31–34]. A descriptive summery of the types of ML models included is depicted in Fig 3, where Supervised ML models, such as random forest and logistic regression were frequently reported, while the reinforcement and unsupervised ML models were less reported.

**Figure 3.**
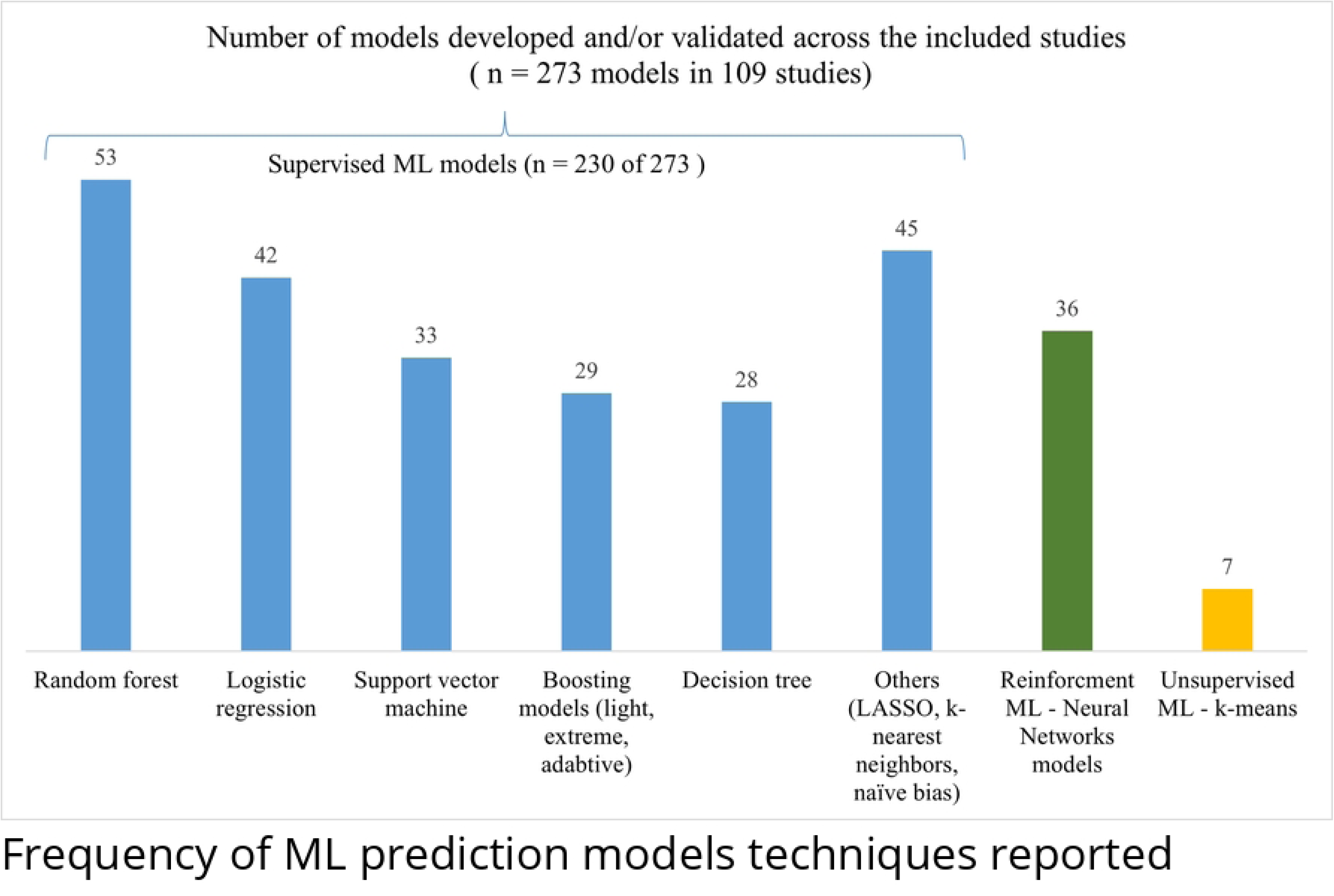
Number of models developed and/or validated across studies

**Figure 4.**
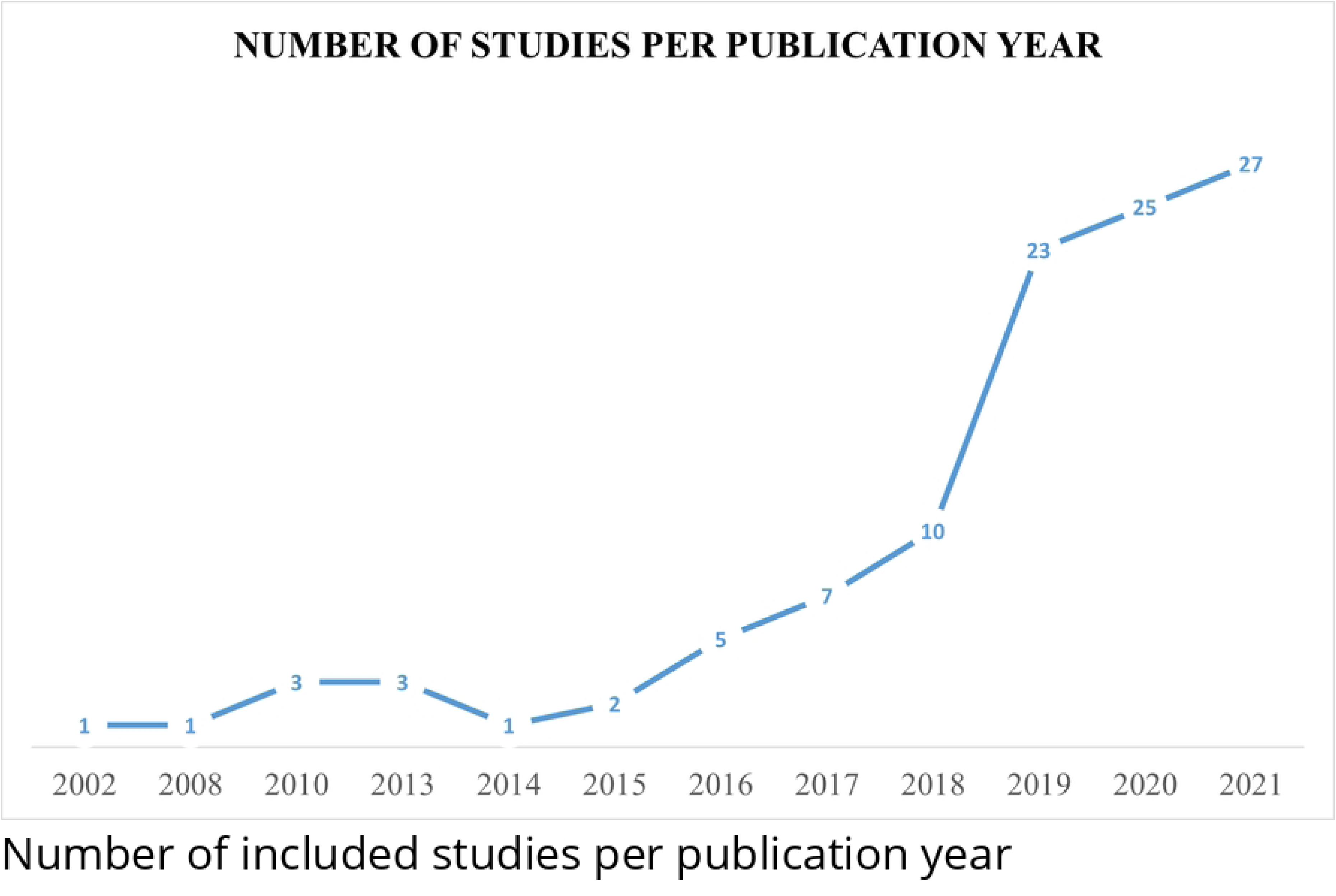
Percentage presentation of the results of PROBAST of the two components: risk of bias (4 domains: Participants, predictors, outcome, and analysis) and concern of applicability (3 domains: Participants, predictors, and outcome)

A few studies (n=10) compared the performance of the developed ML models to other standard reference techniques that were based on classical statistics, such as classical logistic and Cox regression. In seven studies of them, it was reported that ML models outperformed the classical statistics, providing better insights to discover new associations [29,35–40]. The other studies (n=3) reported either similar [41] or lower performance of ML to classical models.[42][43].

Models’ developing attributes, such as features selection and handling of missing data were reported in 68 and 38 studies (of 109), respectively. Models’ internal validation using n-fold cross validation and random splitting of the datasets, either one of them or both in the same study, were reported in 90% and 80% of the included studies, respectively. Broader external validation scale was reported in 25 studies in one different setting or more, such as temporal, geographical, or using different population sample validation. On the other hand, models’ performance measures of discrimination ability using the area under the receiver operating characteristic curve (AUROC) were reported in 62 studies, where results of these measures range from zero for no discrimination ability to ten for the best ability. One study reported the performance measures using decision analysis curve [116].

Tables 1, 2, and 3 present an overview of the included studies characteristics based on the development and validation stage of the models, grouped according to the ICD-10 classification, and ordered alphabetically within each classification. For each study, study design and the objective of the ML prediction model (incident diagnostic, prevalent diagnostic, or prognostic) were provided. Furthermore, the dataset used in each study was reported based on the national location of the dataset and the health care level source being exclusive if only from a PHC data source or linked if PHC data was reported to be linked to other health care data sources, such as secondary or tertiary health care. Last three columns presented the number of the dataset’s population, the timeframe of the data extracted from the dataset used, and the health condition addressed. Nevertheless, in Tables 2 and 3, each study was presented in multiple rows based on the number of the locations used to validate the ML models. An additional panel summary of all the included studies is presented as S5 Appendix.

### Health conditions

Out of the 22 classifications of the ICD-10 version 2019, 11 classifications were addressed in the included studies. Frequently reported classifications were the endocrine, nutritional, and metabolic diseases classification (ICD-10: E) (n= 27 studies of 109, 24.7%), circulatory system diseases (ICD-10: I) (n= 23, 21.1%), and the mental and behavioral disorders classification (ICD-10: F) (n= 22, 20.1%). To a lesser extent, diseases of the respiratory system classifications (ICD-10: J) and neoplasms (ICD-10: C) were addressed in (n=12, 11% and n= 8, 7.3% respectively). 35.9% of the included studies represent other health conditions from the remaining six ICD-10 classifications included. The health conditions addressed are depicted in Tables 1, 2, and 3 and S5 Appendix summary panel.

#### Endocrine, nutritional and metabolic diseases (E00-E90)

In 27 studies addressing this classification [29,31,67–75,116,33,117,118,129– 133,38,40,42,63–66], populations involved were from 12 countries, mainly the US (41.9%). The studies were published since 2008 with the highest number of studies in 2019 (38.7%). 81% of the included studies reported the development and/or training of the proposed models using exclusive primary health care data of a total number of 4.2 million participants. Data was extracted from different data sources over six months up to over almost 23 years. Four health conditions were identified in this ICD-10 classification, namely diabetes mellitus (E10, E11) with/without complications (n= 21), familial hypercholesterolemia (E78) (n= 3), children obesity (E66) (n= 2), and primary aldosteronism (E26) (n= 1). Incident diagnostic prediction was the most frequently reported outcome (42%). Prevalent diagnostic and prognostic prediction were 32% and 26% respectively. Diabetic retinopathy was the most common complication tackled (n=5 of 21 related diabetes mellitus studies), with using not routine primary health care investigations, such as fundoscopy that is used by the secondary health care. Diabetic foot identification was tackled in only one study using only the free text written by the physicians in the form of clinical notes as a predictor [31]. Two studies investigated prognostic predictive modelling of the short- and long-term levels of HbA1c after insulin treatment [72, 116].

#### Mental and behavioral disorder (F00 – F99)

In 22 studies of this ICD-10 classifications addressing six health conditions [28,45,89– 97,119,81,120,121,82–88], the involved population were from eight countries, mainly the US and the UK (n=14). These studies were published since 2013 with the highest number of studies in 2020 (44.4%). Data was extracted from different data sources with varying periods of health records follow up, from one year to almost 28 years. Dementia/Alzheimer’s disease (F00) was addressed in 13 studies, of which one study predicted it within the progression of mental cognitive impairment [85], while another study predicted hospitalization risk [96]. Major depressive disorder (F32) (n= 3) which a study predicted its prognosis within two years and suggested considering the severity of the baseline symptoms for depression prediction [81]. A study claimed to be the first to predict first episode psychosis (F29) and suggested that considerable proportion of the most predictive features were not of a psychiatric nature [45]. A study predicted anxiety (F41) in cancer survivors seeking care in PHC and suggested that fatigue and insomnia were the most important predictors [86]. Lastly, a study used PHC data to predict any mental disorder using different ML modes, claimed that the potentially successful prediction was the best before 180 days of real diagnosis [93].

#### Circulatory and respiratory health conditions (I00-I99 and J00-J99)

In 35 studies addressing these two ICD-10 classifications, populations involved were from 11 countries, mainly the US and the UK. All the included studies were published since 2010 with the highest number of studies in both groups in 2020 (30.8%). Data was extracted from the different data sources over highly variable period from one month to almost 23 years of longitudinal data.

Six circulatory health conditions were identified in 23 studies [27,34,51– 60,35,61,127,128,37,43,46–50]. These conditions were hypertension (I10-I15) (n= 5), heart failure (I50) (n= 5), atrial fibrillation (I48) (n= 2), stroke (I64) (n= 2), atherosclerosis (I70) (n=1), myocardial infarction (I21) (n=1), and any cardiovascular event or disease (n=7). Variable conclusions were reported across these studies. For example, a study reported that variations in the importance of different risk factors depend on the modelling technique [35]. Another study reported that ignoring censoring substantially underestimated risk of cardiovascular disease [43]. Also, systolic blood pressure could be used as a predictor of hospitalization or mortality [59]. Lastly, predictions levels increase after two years and 4000 patients as requirement to predict incident HF cases, and variability of the best performing model could depend on the method of handling the missed data [53].

Five respiratory health conditions were identified in 12 studies [30,32,114,115,41,107– 113]. Chronic obstructive pulmonary disease (COPD) (J40) (n= 5) studies were prognostic predictive studies tackling mortality and hospitalization. Asthma (J45) (n= 2) studies identified known undocumented cases and predicted exacerbation prognosis. COVID-19 (U07) incident cases were predicted within routine PHC visits in one study [111]. With employing results of polychain polymerase reaction (PCR) as predictors, a tree classification approach was reported to be potentially useful in detecting the existence of COVID-19 infection [111]. Contact with a previously infected person was reported as the key factor linked to the development of COVID-19, with recommendation to early detect and isolate the contacts [111].

#### Other health conditions

Eight studies addressed three neoplasms, namely colorectal cancer (CRC) (C18) (n= 6), lung cancer (C34) (n= 1), and pancreatic cancer (C25) (n= 1). Four studies addressed the same incidence prediction model known as ColonFlag (previously MeScore) to identify CRC cases [125,134,136,137]. Each study predicted incident cases within different time windows before diagnosis; over three months to two years with relative high discrimination ability of the proposed model across the four studies. This ColonFlag model was reported as ‘well-performing’ when used on CRC cases detected at early asymptomatic (often nonanemic), localized stages, as well as when limited to complete blood picture data collected around a year before diagnosis [125].

Three health conditions affecting the nervous system were addressed [104–106], one of which predicted mortality four years before and after diagnosis of epilepsy (G40) with an acceptable performance for identifying those at high risk of early premature mortality [105]. Another study predicted a rare neurodegenerative disease, progressive supranuclear palsy (G23), identified two previously unknown clinical features as predictors associated with the pre-diagnostic stage of this disease [106].

Regarding musculoskeletal and connective tissue disorders [98–100,122–124], back pain (M54) prognosis within PHC settings could be predicted through focusing on patient function-related predictors more than on resolving pain [99]. A study revealed that models can be created using only data from medical records and had prediction values of 70-80% for identifying persons who are at risk of acquiring ankylosing spondylitis (M45) [100]. Two digestive health conditions were addressed in two studies [36, 62], which were inflammatory bowel diseases (K50-K52), including Crohn’s disease and ulcerative colitis, and peptic ulcers (K27)/gastroesophageal reflux (K21). Two studies addressed the chronic kidney disease (N18) [28, 79], one of them was an incident diagnostic and the other predicted hospitalization and steroid use within 6 months and one year. Three studies tackled suicidality (X60-X84) [76–78], one of which predicted incidence of suicide and reported that PHC records were of little indication of severity [76]. Lastly, one study addressed preeclampsia (O14) with additional reporting of a systematic review of this disease across different health care sectors [126].

### Quality assessment

Addressing the included studies using the PROBAST tool revealed that 90.8% (n=99 of 109) of the included studies were of high and unclear risk of bias, as depicted in Fig 5. Analysis domain was the main source of bias, because of underreporting. Additionally, the studies of potential low risk of bias were downgraded from high risk due to the of lack of external validation of the proposed models (n=20). Only a few studies (n= 11) were reported in accordance with transparent reporting of a multivariable prediction model for individual prognosis or diagnosis (TRIPOD) guidelines [138]. Concern of applicability of the addressed models in the PHC of 72 (66%) studies was low. The main source of this concern is the dependence of the predictive models on not-routine PHC data.

Most of the included studies (n= 101 of 109, 92.7%) were published as peer-reviewed publications in biomedical (e.g., PLOS ONE, n= 8) and technical journals (e.g., IEEE, n= 3). Eight studies were preprint and abstracts. National research institutes and universities were the most frequently reported funding support. Most of the studies reported that the funding supporters were not involved in the process or results of the published work. Nevertheless, some studies were supported by industrial companies without clarifying the role of the funding body.

## Discussion

ML prediction models could have an immense potential to augment health practice and clinical decision making in various health sectors. Our systematic review provides an outline and summary of the health conditions tackled through ML prediction models using PHC data.

42 health conditions across 109 observational studies were identified. 76.6% of the included studies were diagnostic, while 23.4% were predictive of complications, hospitalization, or morbidity. Alzheimer’s disease, diabetes mellitus, heart failure, colorectal cancer, and chronic obstructive pulmonary diseases were the most frequently targeted health conditions. Less attention was directed to the other reported diseases, such as asthma, children obesity, and dyspepsia.

In the context of PHC, detection and management of evitable and controllable chronic health conditions, such as diabetes mellitus are part of the most vital role of this health care settings [3]. On the other hand, misdiagnosis of diseases can result in abandoned symptoms, ineffective treatment, and preventable deaths [3]. Despite of the early stage of the ML prediction models of such health conditions in PHC [139], primary care setting have gained more attention in many countries [11], similar to our findings. Furthermore, predicting undocumented cases and rare diseases with potential good performance was also reported. Nevertheless, it is suggested to investigating the prediction of other diseases incidence and progression among the health care providers, researchers, and models’ developers.

Health conditions diagnostic and prognostic predictions were performed using 273 ML models mainly of supervised learning technique. The models within 77% of the included studies were trained and/or internally validated without evaluating their generalizability. The other 33% of studies present those conducted internal and/or external validation. Despite relatively good performance reported across the included studies, their clinical implication is limited, and further investigations are needed. Furthermore, lack of reporting guidelines usage and overall risk of bias assessment of high to unclear raise concerns about the potential disadvantages of such models.

Technical biases could influence the clinical practice. When a model is trained on historical data, which supports old practice without adaptation to policy changes, then the model reinforce an outmoded practice [140]. Furthermore, due to bias in the training set, change over time, or application of the system in a different population, a mismatch between the data or environment on which the system is trained and that used in operation may result in an erroneous result [140]. This bias could affect the results of some of the included models being trained with data generated up to 40 years ago. Additionally, lack of reporting the different health systems prevents the proper estimation of the applicability of external validation results. Hence, it is recommended to properly report the development and performance measures attributes of the models under progress in the presence and the future. Additionally, it is encouraged to validate the proposed models within different geographical and temporal settings with proper reporting of the main up-to-date criteria of the health system addressed as well.

The main source of the extracted participants’ data was exclusive primary health care data. Various other sources of data were linked to the PHC data, such as secondary and tertiary health care. These health data represented a total of 24.2 million participants mainly within PHC settings. A large majority of the models’ development and/or validation was conducted in the US and the UK (58.1%) with a noticeable rise since 2018.

Despite the fact that big data generated through the health records is a strong fit for ML tools, the coding system itself does not universally follow the same criteria for diseases [141]. Furthermore, PHC has no standardized definition globally with a wide variability of the services provided. Hence, different health system and terminology of diseases and symptoms across the world could limit the consistency of the models’ performance [141]. Additionally, uncoded free-text clinical notes and the lack of proper coding, such as using (‘race’ and ‘ethnicity’) and (‘suicide’ and ‘suicide attempts’) to be documented as a single input, affect the predictive power of the models [142]. Other drawbacks reported, similar to our findings, were underrepresentation of healthy persons and retrospective temporal dimension of the extracted predictors [142]. Therefore, routine care data collected according to a documentation system might not fully match the proposed questions with the models’ developed. Additionally, misclassification bias and incomplete health records represented a major limitation, as reported in the included studies. Even with proper classification, certain diseases require confirmatory diagnosis using higher care services, such as magnetic resonance imaging (MRI) [143], which is missing from PHC. Therefore, it would be advisable that models’ developers propose solutions for the digital documentation systems, when possible, based on the addressed health condition to overcome the limitations faced with discussing these solutions’ benefits and applicability. With that approach, more evidence-based literature would be available for the stakeholders to implement further enhancements.

On the other hand, the unequal distribution of papers across countries could be related partially to the low publication rate in the low-income countries or lack of proper big data documentation systems. However, this justification does not clarify the reason of the unequal distribution of publication among the middle- and high-income countries. Hence, the transition from using the conventional medical records to integrating the predictive models in PHC is far from simple and necessitates specialized processing techniques. Furthermore, solid technical infrastructure as well as strong academic and governmental support are essential for promoting and supporting long-term and broad-reaching PHC data gathering efforts [142, 144]).

Lastly, based on the high variability of the structure and reporting styles identified across the studies, i.e., medical versus engineering point of view and style, it is recommended to augment the participation of health professionals through the development process of the health related predictive models to critically evaluate, assess, adopt, and challenge the validation of the models within practices, given the increasing popularity of digitally connected health systems [5]. Furthermore, ML engineers must be aware of the unintended implications of their algorithms and ensure that they are created with the global and local communities in mind [145]. Hence, it is advisable to obtain an efficient cooperation between ML developers and the health care professionals to provide new insights for tackling the potential biases. Additionally, it is suggested to integrate the basic understanding of ML concepts and techniques among the under- and post-graduate education programs.

### Strength and limitations

Our review was conducted following a predesigned comprehensive protocol [21]. We identified the health conditions targeted within primary care settings as an encompassing of literature and identifying the gaps needed to be tackled. However, the main limitation of our review’s quality of evidence, first, is the reviewing of observational studies that mostly lacked external validation of the proposed models. Second, regarding our search strategy, some studies could have been missed if they exclusively used ‘big data’, ‘statistical modelling’, ‘statistical learning’ or similar terms instead of our search string as noted in [146]. Third, limiting our scope to the clinical health conditions resulted in excluding other conditions that could be reported and predicted within the PHC, such as domestic violence and drug abuse [3]. Fourth, guiding our work using ICD-10 might lead to excluding potentially relevant studies, such as a study that used frailty as a medical syndrome [147]. Lastly, we neither extract thoroughly the performance measures of each study nor conduct a meta-analysis, because of the broad heterogeneity across studies. In the future, we plan to update our review - considering this noticeable rise of the PHC ML studies – while also modifying our methodology to reduce the identified limitations. Additionally, we plan to use the new under-progress specific ML guidelines TRIPOD-AI and PROBAST-AI when published to strengthen quality and reporting of our findings [148].

In conclusion, ML prediction models within PHC is gaining traction. Further studies are needed, especially those with prospective designs and more representative samples. Working among multi-discipline teams to tackle ML in primary care increases the trust of the models and their implementations with further consideration of improving quality of development and reporting of the ML predictive models. More research is required to continue to fill the gaps in knowledge surrounding the emergence of PHC data.

## Data Availability

All relevant data are within the manuscript and its Supporting Information files.

## Acknowledgment

Dr. Marcos André Gonçalves, PhD and his colleague Bruna Zanotto, MSc. provided their feedback on the project’s primary draft. Dr. phil. Luana Fiengo Tanaka retrieved the inaccessible studies.

## Supporting information

S1 Appendix Search strategy

S2 List of excluded studies with reasons (n = 58)

S3 Figure Countries under study with total number of studies per country

S4 Figure of studies designs

S5 Appendix Panel of the included studies’ (n =109) characteristics S6 Prisma Checklist

## Notes

### Competing Interest Statement

The authors have declared no competing interest.

### Clinical Protocols

https://www.crd.york.ac.uk/prospero/display_record.php?RecordID=264582

### Funding Statement

The authors received no specific funding for this work.

